# ARID1A immunohistochemical expression is associated with metachronous colorectal polyp risk: Evidence from integrated genomic and transcriptomic analyses

**DOI:** 10.1101/2025.05.03.25326857

**Authors:** Aula Ammar, Amna Matly, Kimiya Kabiri Arani, Scott Murray, Mayuri Hendricks, Alexander Winton, Natalie Fisher, Noori Maka, Gerard P Lynch, Jennifer Hay, Tomoko Iwata, Colin W Steele, Mark S Johnstone, Philip D Dunne, Stephen T McSorley, Joanne Edwards

**Affiliations:** School of Cancer Sciences, University of Glasgow, UK; National Health Service Greater Glasgow and Clyde (NHSGGC), Glasgow, UK; Patrick G Johnston Centre for Cancer Research, School of Medicine, Dentistry and Biomedical Sciences, Belfast, UK; Cancer Research UK Scotland Institute, Glasgow, UK; School of Medicine, Dentistry and Nursing, University of Glasgow, UK

**Keywords:** ARID1A, colorectal cancer, screening, early detection, immunohistochemistry

## Abstract

Current post-polypectomy surveillance strategies rely on morphological features and fail to capture the biological heterogeneity that underlies early colorectal neoplasia. While the SWI/SNF subunit ARID1A is frequently altered in colorectal cancer, its clinical utility and biological role during the pre-malignant stage remain unclear.

We evaluated ARID1A protein expression via immunohistochemistry (IHC) in colorectal polyp tissue microarrays (TMAs, n=1184). To define the underlying biology, we integrated somatic mutation profiling, bulk RNA sequencing and multiplex immunofluorescence for immune context (CD3, CD8, FOXP3, and CD68).

High ARID1A expression was independently associated with a significantly increased risk of metachronous lesions (*p*<0.001), consistently stratifying patients otherwise classified as low risk by conventional criteria (e.g., single polyps). Somatic ARID1A mutations correlated with reduced protein expression (*p*<0.001), supporting a genetic basis for these divergent phenotypes. Transcriptomic profiling revealed that high-ARID1A polyps were driven by Myc-target pathways, high cell proliferation, transcriptional activation and a pronounced regenerative stem-cell signature, alongside increased FOXP3+ regulatory T-cell infiltration. Conversely, ARID1A-low lesions were characterised by transcriptional repression, cell-differentiation pathways, and increased CD8+ T-cell infiltration.

Tissue ARID1A expression captures the underlying pathogenic heterogeneity in pre-malignant colorectal disease, linking distinct genomic, transcriptomic and immune profiles directly to clinical behaviour. Assessing ARID1A status provides a powerful, biologically informed framework for improving post-polypectomy risk stratification beyond conventional clinicopathological criteria.

## Introduction

Bowel surveillance guidelines following polypectomy rely on basic pathological features to assess metachronous polyp or colorectal cancer (CRC) risk. The main factors in different surveillance guidelines include polyp count, histological subtypes and dysplasia grade (1–3). However, in our recently published audit of a large UK screening cohort, 48% of patients classified as low risk according to the British Society of Gastroenterology (BSG) 2020 surveillance guidelines subsequently developed metachronous lesions, whereas 39% of those classified as high risk did not (4). Therefore, a significant limitation of current stratification methods exists in identifying future lesions, suggesting that the inclusion of molecular markers could enhance predictive accuracy. Despite growing evidence of their potential utility, molecular features are remarkably absent from current surveillance guidelines.

Genetic studies have linked multiple mutations to the risk of metachronous or advanced adenomas (reviewed in (5)); however, the mutational landscape of pre-malignant colonic polyps remains poorly defined. A recent single-cell study profiled 21 pre-cancerous colonic lesions, covering both adenomatous and serrated histology, and compared the mutation rates across tubular, tubulovillous, and serrated subtypes (6). However, a lack of longitudinal follow-up data limits its ability to associate specific mutations with future risk of metachronous disease. Therefore, there remains a need to identify molecular biomarkers that not only characterise pre-malignant lesions, but also predict subsequent metachronous neoplasia.

Among the potential candidate biomarkers, ARID1A has attracted considerable interest because it encodes a key component of the SWI/SNF chromatin-remodelling complex and is widely recognised as a tumour suppressor that is frequently altered across several malignancies, including colorectal cancer (7). However, accumulating evidence suggests that the biological consequences of altered ARID1A activity are highly context dependent. In normal colonic epithelium, ARID1A haploinsufficiency reshapes epithelial fitness and promotes age-dependent expansion of mutant clones (8). On the other hand, *ARID1A*-mutant epithelium appears resistant to chemical tumour initiation, as mice harbouring *ARID1A* mutations failed to develop the increased tumour burden observed with other priming mutations following ENU mutagenesis (9).

Further evidence suggests that the biological effects associated with *ARID1A* alterations depend on a broader genetic context. In murine intestinal models, co-mutation of *ARID1A* and *APC* has been associated with enhanced epithelial differentiation and improved survival outcomes (10). In contrast, concurrent loss of *ARID1A* with *TP53* or *PTEN* promotes aggressive tumour phenotypes in endometrial and pancreatic cancer models (11, 12) indicating that the functional consequences of ARID1A deficiency are strongly influenced by coexisting genetic alterations.

Mechanistically, ARID1A loss may induce telomere cohesion defects, resulting in cell-cycle arrest or mitotic death in cells with substantial chromosomal abnormalities, thereby preferentially allowing the survival of clones with lower levels of chromosomal instability (13). Consistent with this observation, some *ARID1A*-mutant colorectal tumours exhibit fewer large-scale copy number alterations than their wild-type counterparts (13). Together, these findings suggest that ARID1A influences early clonal dynamics and genomic stability rather than functioning as a conventional tumour suppressor.

The prognostic significance of ARID1A expression remains similarly complex and appears to be highly tissue- and context-dependent. Studies evaluating ARID1A expression have reported associations with both adverse and favourable clinical outcomes across different tumour types and disease stages (14–18). These conflicting observations suggest that ARID1A may not function as a universal prognostic marker; rather, its clinical relevance is likely influenced by the tumour context, disease stage and coexisting molecular alterations. Understanding how differences in ARID1A expression are related to the biology of pre- malignant colorectal lesions remains an important unmet need.

Despite growing evidence that ARID1A plays an important role in epithelial homeostasis and early colorectal tumorigenesis, its prognostic significance in pre-malignant colorectal polyps remains unclear. In this study, we used a well-annotated cohort of patients with large precancerous colorectal polyps to evaluate the prognostic value of ARID1A protein expression. By integrating immunohistochemistry, mutational profiling and transcriptomic analyses, we demonstrated that ARID1A within the index polyp independently predicts a higher risk of subsequent metachronous disease, providing a potential molecular refinement of current colorectal surveillance strategies.

## METHODS

### Patient cohort and tissue microarray (TMA) construction

TMAs from formalin-fixed paraffin-embedded (FFPE) index colonic polyps representing the **IN**tegrated te**C**hnologies for **I**mproved **P**olyp Surveillanc**E** (INCISE) cohort (4) were tested for ARID1A expression. These polyps were removed at bowel screening colonoscopy, and all patients underwent at least one surveillance procedure up to 6 years after the initial colonoscopy. This study excluded polyps associated with inflammatory bowel disease (IBD), inflammatory polyps, familial adenomatous polyposis (FAP), and hyperplastic rectal polyps (4). The term “index polyp” represents the largest (≥1cm) and/or most advanced polyp from patients aged 50-74 yrs. For TMA construction, the largest/most advanced polyps were considered most clinically significant according to the BSG2020 guidelines (1) therefore TMA mainly included large polyps ≥1 cm. Four 0.6-mm cores per patient that captured tissue heterogeneity were included. A total of 1201 patients were initially included in the TMA. Seventeen cases were excluded from the analysis due to tissue loss or folding during the staining procedure, resulting in a final cohort of 1184 patients. The clinicopathological characteristics of the study cohort are summarised in Table 1.

**Table 1.** Patient characteristics and Chi-squared test results of epithelial ARID1A histoscores with clinicopathological criteria.

|  | Total numbers | ARID1A (n=1184) |  |  |
| --- | --- | --- | --- | --- |
|  | N(%) | Low N(%) | High N(%) | p value |
| Sex |  |  |  |  |
| Male | 858(72.5) | 155(18.1) | 703(81.9) | 0.272 |
| Female | 326(27.5) | 68(20.9) | 258(79.1) |  |
| Age (median) |  |  |  |  |
| ≤63 y | 630(53.2) | 129(20.5) | 501 (79.5) | 0.123 |
| >63y | 554(46.8) | 94(17.0) | 460 (83.0) |  |
| Site of most advanced polyp |  |  |  |  |
| Right colon | 165(13.9) | 16 (9.7) | 149(90.3) | <0.001 |
| Left colon | 861(72.7) | 166(19.3) | 695 (80.7) |  |
| Rectum | 155(13.1) | 41(26.5) | 114 (73.5) |  |
| missing | 3(0.3) |  |  |  |
| Histology |  |  |  |  |
| T | 504(42.6) | 92 (18.3) | 412 (81.7) | 0.567 |
| TV | 588(49.7) | 118(20.1) | 470 (79.9) |  |
| V | 71(6.0) | 10(14.1) | 61 (85.9) |  |
| Serrated features | 21(1.8) | 3(14.3) | 18 (85.7) |  |
| Dysplasia grade |  |  |  |  |
| Low grade | 996(84.1) | 189(19.0) | 807(81.0) | 0.774 |
| High grade | 188(15.9) | 34(18.1) | 154(81.9) |  |
| Number of polyps |  |  |  |  |
| 1 | 380(32.1) | 75 (19.7) | 305(80.3) | 0.861 |
| 2-4 | 636(53.7) | 117(18.4) | 519(81.6) |  |
| ≥5 | 168(14.2) | 31(18.5) | 137(81.5) |  |
| Future polyp detection |  |  |  |  |
| No | 518 (43.8) | 123(23.7) | 395 (76.3) | <0.001 |
| Yes | 666 (56.3) | 100(15.0) | 566 (85.0) |  |
| Future advanced lesion status |  |  |  |  |
| No future polyp | 518(43.8) | 123(23.7) | 395 (76.3) | <0.001 |
| Non-advanced | 465(39.3) | 67(14.4) | 398 (85.6) |  |
| Advanced | 201(17.0) | 33(16.4) | 168 (83.6) |  |
| BSG2020 score |  |  |  |  |
| Low | 385(32.5) | 76(19.7) | 309(80.3) | 0.580 |
| High | 799(67.5) | 147(18.4) | 652(81.6) |  |

The median follow-up period was 36 months (range: 6-72 months). Patients were right censored at 6 years. Ethical approval and patient consent was granted through the INCISE project (GSH/20/CO/002) (4).

### Immunohistochemistry (IHC) and multiplex immunofluorescence (mIF)

ARID1A antibody specificity was confirmed prior to immunohistochemistry using Western blotting (supplementary methods). IHC was used to study protein expression in cell pellets and polyp tissues. Ultravision Quanto (UVQ) detection system (UVQ, Epredia) was used for ARID1A (Cell Signaling, #12354, 1:1500) and CDK4 (Cell Signaling, # D9G3E, 1:100) and counterstained with haematoxylin. Staining with c-Myc antibody (Abcam, #ab32072, 1:800) and Ki67 (Agilent, #M7240, 1:100) was performed using a Leica Bond Rx autostainer and Refine Kit (Leica Biosystems). mIF staining of TMA sections was performed using UVQ kit with primary antibodies against CD3 (Leica, #NCL-CD3-565), FOXP3 (Abcam, #ab20034), CD8 (Agilent, #M7103), CD68 (Agilent, #M0876), αSMA (Agilent, #M0851) and pan- Cytokeratin (Theromfisher, #MS-343-P) as described previously (19). The signal was visualised using the OPALTM reagents and scanned using a PhenoImager HT (AKOYA Biosciences) at 20x magnification. Full protocols’ details are provided in the supplementary methods. QuPath (v0.5.0) (20) was used to analyse IHC and mIF images as detailed in the supplementary methods. IHC histoscores (H-scores) were calculated by multiplying the percentage area scoring positive by the respective intensity using the following formula: (% of cells stained weak × 1) + (% of cells stained moderate × 2) + (% of cells stained strong × 3) (range 0-300) (21).

### Genomic DNA extraction, sequencing and somatic mutational analysis

DNA was extracted from 10-µm tissue sections (n=623). Targeted sequencing was performed using the Agilent SureSelect XT2 HS2 Cancer Plus panel on the NovaSeq 6000. Next-generation sequencing data processing was performed using the HOLMES pipeline v1.3.1. Somatic mutation analysis was performed using Maftools (v2.18.0) in R. ARID1A mutations were visualised with Mutation Mapper on cBioPortal (22, 23) and compared to the OncoKB list (24) to assess pathogenic/oncogenic mutations (accession date Dec 2024). Statistical significance was set at *padj*<0.05.

### Transcriptomics analysis

Templated Oligo assay with Sequencing (TempO-Seq) readout (Biospyder Technologies, USA) whole transcriptome v2.0 panel profiling was performed on 4µm full sections (25). Quality control, preprocessing and analysis packages are described in supplementary materials. in total, 234 samples (117 low and 117 high ARID1A) were included in this study.

### Statistical analysis

ARID1A histoscores were divided into low and high expression groups using an objective, data-driven cut-off generated with the R package “survminer”. This method systematically evaluates all possible thresholds and selects the value that most effectively discriminates patient outcomes (e.g. survival), avoiding subjective or arbitrary categorisation. IHC results were analysed by correlating ARID1A expression (low/high) with clinicopathological and mutational data using Chi-squared tests. Kaplan-Meier survival curves were plotted, and group differences were assessed using log-rank tests. Cox-regression multivariate analysis, adjusted for confounding clinical/pathological factors, was performed. Statistical significance was set at *p*<0.05. Both IBM SPSS Statistics (ver28) and in R were used for data processing and analysis.

## Results

### ARID1A expression in colonic polyp tissue and its association with clinicopathological criteria

Nuclear ARID1A was expressed in epithelial and lamina propria cells of the normal colon. In adenomatous crypts, epithelial cells showed heterogeneous staining intensities ranging from weak to strong, as well as a mosaic staining pattern and partial or complete loss of expression Figure 1, A-F). Following Qupath quantification of histoscore (Figure 1-G), the median H-scores were 183.6 (range 16.0-264.93). The cutpoint of 147.49 was mathematically generated using the survminer package in R (Figure 1-H). Supplementary Figure 1-A shows the specificity check of ARID1A antibody using Western blotting and cell pellet staining.

**Figure 1.**
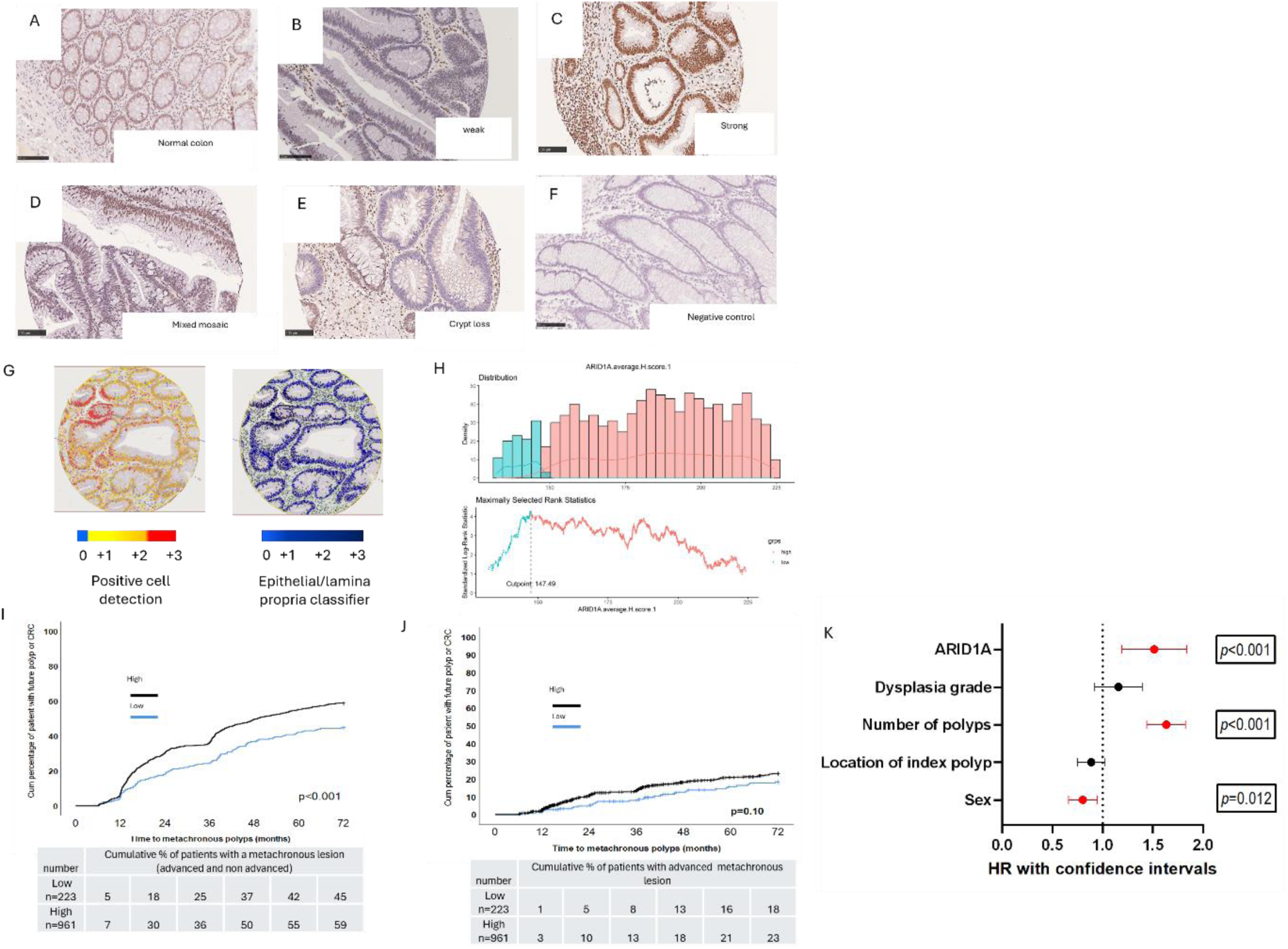
IHC of ARID1A in colonic pre-malignant polyps predictive significance of ARID1A for metachronous colorectal lesion. **(A)** A representative example of ARID1A staining in normal colon. (B-E) Weak, strong, mosaic and crypt-loss expression ARID1A expression in polyps **(F)** A representative example of no primary negative control. **(G)** QuPath positive cell detection with different intensities and adenoma/lamina propria classifiers. **(H) G**eneration of the ARID1A cutpoint using the survminer package in R in the training set showing a cutpoint of 147.49.**(I)** Time to event curves for ARID1A high/low groups with regards to metachronous lesions over 72 months of follow-up. ARID1A expression was significantly associated with future polyp outcomes (Log-rank test, *p*<0.001). Life tables showing percentages of patients developing the outcome. **(J)** Time to event curves for ARID1A high/low groups with regards to advanced metachronous lesions status (Log-rank test, *p*=0.10). Life tables showing percentages of patients developing an advanced future lesion. (**K**) Cox-regression multivariate analysis for ARID1A and other confounding factors. ARID1A, number or polyps and sex were significant indicators of outcome in the model. *p* values ≤0.05 were considered statistically significant.

Chi-squared tests revealed significant associations between the ARID1A histoscore groups and polyp location (i.e., right colon, left colon or rectum; *p*<0.002), future polyp status (*p*<0.001) and advanced future polyp status (*p*<0.001) (Table 1). Time to event analysis demonstrated that high ARID1A histoscores were significantly associated with future polyp detection (Figure 1-I) with univariate analysis HR ratio of 1.517 (confidence interval (CI): 1.226-1.877, *p*<0.001). The predictive value of ARID1A histoscores for *advanced* future polyp status was also investigated (Figure 1-J). While the low- and high-ARID1A groups displayed an observable separation, the association was not statistically significant (*p*=0.10). These findings suggest that ARID1A expression may serve as a predictor of future polyp occurrence rather than specifically identifying patients at risk of developing metachronous polyps with advanced features. The median value of ARID1A-Histoscores was also tested as an alternative cutpoint and similar significant results were observed between ARID1A expression and the location of the index polyp, histology, metachronous polyp occurrence and advanced future polyp status (Supplementary Table 1).

### ARID1A is predictive of metachronous polyp outcome in multivariate analysis

Multivariate Cox regression was performed to assess ARID1A expression while adjusting for confounding factors including sex, index polyp location, number of polyps, and dysplasia grade (tested in univariate analysis with significant p values for sex, location, and number of polyps as shown in supplementary figure 1-B). This demonstrated that ARID1A-histoscore was an independent predictor of metachronous polyp risk (hazard ratio (HR)=1.491 (CI:1.204-1.847), *p*<0.001; Figure 1-K). ARID1A demonstrated 84.98% sensitivity and 23.75% specificity and accuracy of 58.19%. The high sensitivity of ARID1A expression suggests potential utility as an additional screening marker to identify patients who may warrant closer surveillance or additional molecular testing. When the median cutpoint was used, ARID1A was also an independent predictive factor with HR=1.225 (CI:1.050-1.29) and *p*=0.010.

### ARID1A predicts metachronous lesions development in lower risk patients

ARID1A predictive significance was further analysed in subgroups stratified by the number (1, 2-4, or ≥5), location of index polyps and dysplasia grade to explore whether ARID1A could improve risk assessment in patients considered to have lower clinical risk. ARID1A showed the strongest predictive value in patients with fewer than five polyps (i.e., groups with 1 polyp or 2-4 polyps; *p*=0.034 and *p*=0.003, respectively; Figure 2, A-D). Analysis of ARID1A predictive value by polyp location showed that patients with index polyps in the left colon and rectum may benefit more from incorporating ARID1A as a predictive marker (*p*=0.001 and *p*=0.053, respectively) than those with right-sided polyps (*p*=0.792) (Figure 2, E-H). Similarly, ARID1A high expression was a strong predictive factor for metachronous polyps in patients with LGD polyps (*p*<0.001; Figure 2, I-J).

**Figure 2.**
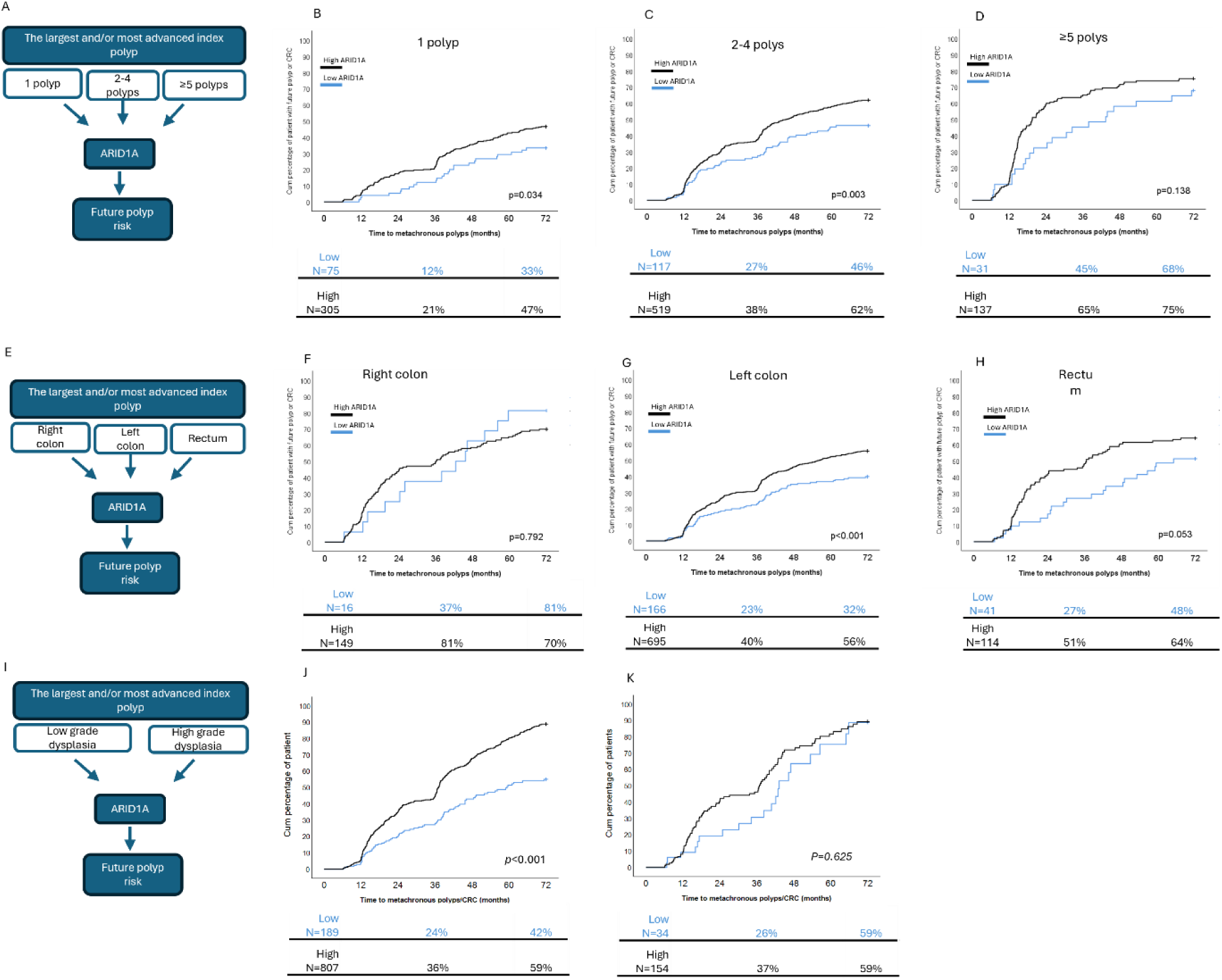
Time to event analysis of ARID1A in subgroups of patients based on number of polyps, location of index polyp and dysplasia grade. (A-D) ARID1A predictive significance in patients’ subgroups based on number of polyps at index colonoscopy (1 polyp, 2-4 polyps and ≥5 polyps) showed good predictive value of ARID1A in patients with 1 or 2-4 polyps. (**E-H)** ARID1A predictive significance in patients’ subgroups based on location of index polyp (right colon, left colon and rectum.) showed good predictive value of ARID1A in patients presented with the index polyp in the left colon or the rectum. (**I-K**) ARID1A predictive significance in low- and high-grade dysplasia groups. Life tables showing percentages of patients developing a metachronous polyp at 3 years and 6 years. *p* values ≤0.05 were considered statistically significant. *p* values ≤0.05 were considered statistically significant.

### Reduced ARID1A protein expression in precancerous polyps is partially explained by *ARID1A* mutations

To determine whether reduced ARID1A protein expression was driven by the underlying mutations, we evaluated *ARID1A* mutation status. *ARID1A* was among the most frequently mutated genes in pre-cancerous colonic polyps (Figure 3-A). Mutational frequencies of *APC*, *KRAS*, *MSH3* and *TP53* were similar across polyps with high and low ARID1A histoscores. However, mutations in *KMT2D* and *ARID1A* were significantly more frequent in the ARID1A-low group (12% vs. 24% and 11% vs. 20%, respectively). To validate the functional impact of genomic alterations on tissue phenotype, we integrated our mutational data with raw IHC histoscores. As demonstrated in Figure 3B&C, polyps with ARID1A or KMT2D somatic mutation exhibited significantly lower median histoscores than non-mutant polyps (*p*<0.001 and *p*=0.021, respectively). This result is concordant with a previous report that methylation-related alterations and *ARID1A* mutations may drive ARID1A protein loss at least in CRC cell line models (26). No other mutations were significantly associated with ARID1A protein levels (data not shown). Mapping *ARID1A* mutations to gene exons revealed no recurrent pathogenic mutations in either our polyp cohort or the OncoKB^TM^ colorectal cancer cohort, which was included as a control (Figure 3-D). Exons 1 and 20 were the most affected exons in the polyps (Figure 3-E). No significant difference in ARID1A protein expression was observed based on exon location (ANOVA *p*=0.089, supplementary figure 2A). The most common substitution was C>T (47.96%) transition (Figure 3-F) which is consistent with age-associated endogenous mutational processes, particularly the spontaneous deamination of 5-methylcytosine, which contribute substantially to the mutational landscape of colorectal neoplasia (27). Truncating mutations were the most common ARID1A alterations (48%) and were associated with significantly reduced ARID1A protein expression (Figure 3- G&H). *ARID1A* mutation was not associated with metachronous polyp/CRC development (Supplementary figure 2B) suggesting that ARID1A protein expression is independent of mutational status and may instead reflect broader transcriptional or epigenetic programs within the polyp.

**Figure 3.**
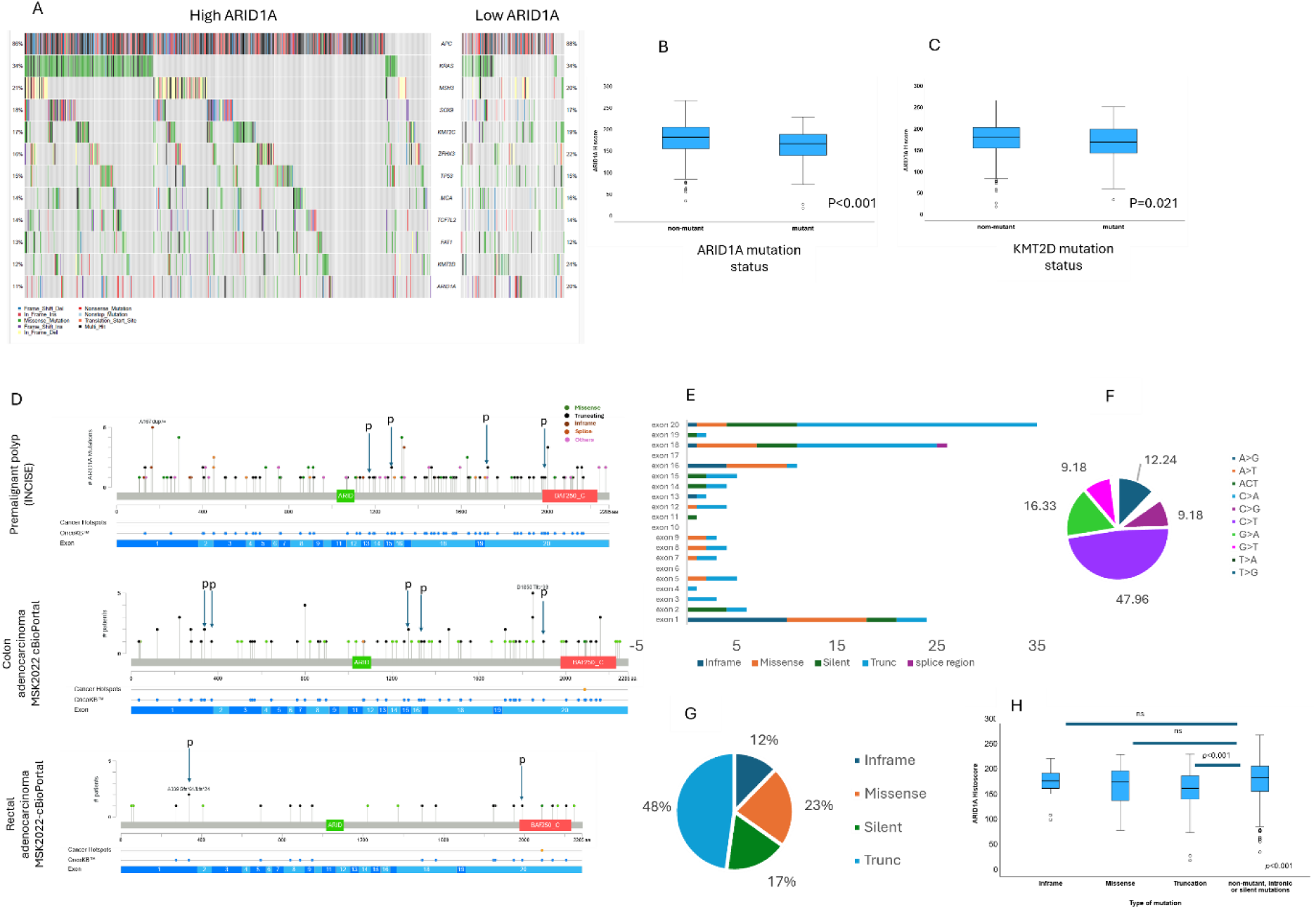
ARID1A mutation and protein expression in pre-malignant colonic polyps: mutational and expression profiling. **(A)** Oncoplot comparing top mutations in colonic polyps between ARID1A high- and low-expression groups. The most frequently mutated genes preceding *ARID1A* were *APC*, *KRAS, MSH3, SOX9, KMT2C, ZFHX3, TP53, MGA, TCF7L2, FAT1* and *KMT2D*. *KMT2D* and *ARID1A* mutation rates were doubled in the ARID1A low-expression group. **(B&C)** Box plots showing a significant decrease in ARID1A protein expression in the presence of *ARID1A* (*p*<0.001) or *KMT2D* (*p*=0.021) mutations. **(D)** Lollipop plots of *ARID1A* mutations in polyps, colon adenocarcinoma and rectal carcinoma across 20 exons. Arrows indicate pathogenic/oncogenic mutations based on the OncoKB™ database. **(E)** Bar graphs of mutation types across *ARID1A* exons, highlighting exon 1, 18 and 20 as the most frequently mutated. **(F)** Percentage of the most common substitution mutations in polyp tissues, with C>T transitions being the most frequent (47.95%). **(G)** Types of *ARID1A* mutations in polyps, showing truncating mutations as the most frequent (48%). **(H)** Box plots showing a significant decrease in ARID1A protein expression in the presence of truncation mutations. *p* values ≤0.05 were considered statistically significant.

### ARID1A expression correlates with Myc pathway activation and proliferative phenotype in colonic polyps

To define the transcriptional landscape associated with ARID1A expression, specimens representing the top and bottom percentiles (n=117 low and n=117 high) were analysed. At the pathway level Myc signalling (Myc_targets_V2 pathway) was enriched in polyp tissues with high ARID1A expression (Figure 4-A), which was confirmed at the single sample level (*p*<0.001; Figure 4-B). When network analysis was performed for the leading-edge genes of the Myc signalling pathway, stronger and more inter-gene correlations were observed among a subset of leading-edge genes in the high ARID1A expression group than in the low ARID1A expression group (Figure 4-C). Notable differences in expression were observed for *Myc* and *CDK4* (Figure 4-C), which are key accelerators of the G1 phase of the cell cycle. To validate this observation, polyp tissue TMAs were stained for Myc, CDK4 and Ki67 proteins (Figure 4D-F). A significant increase in Myc, CDK4 and Ki67 (general proliferation marker) expression was observed in tissues with high ARID1A expression (all *p*<0.001, Figure 4D-F). These results further support the transcriptomic data and validate activation of the Myc pathway in the ARID1A-high expression group, supporting a shift toward a proliferating and self-renewal phenotype in polyps with high ARID1A expression.

**Figure 4.**
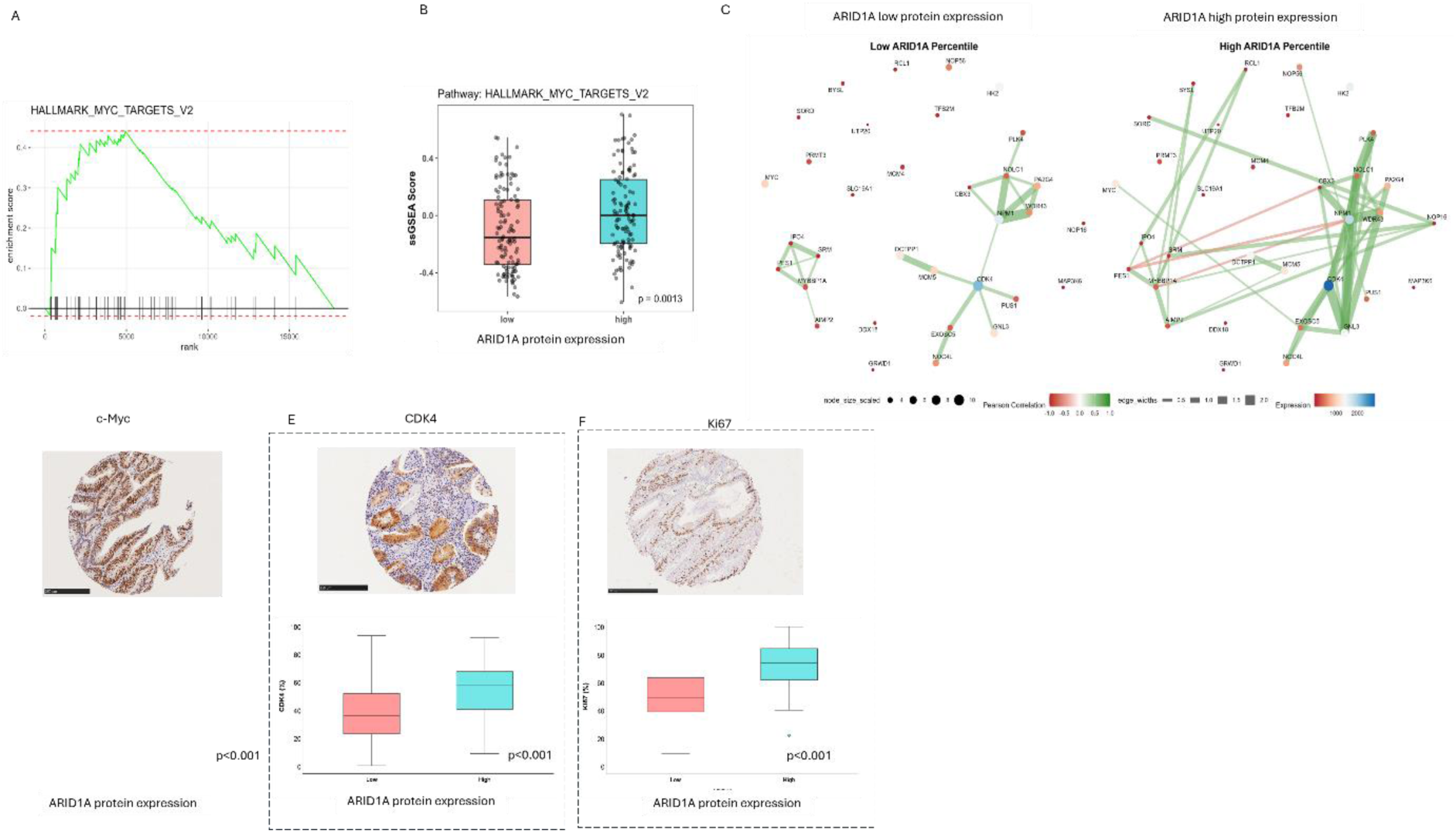
Transcriptomic analysis of polyps grouped by ARID1A expression and validation of protein expression. **(A)** Hallmark pathway analysis using GSEA showing the enrichment plot of Myc-targets-V2 pathway. **(B)** a comparison of ssGSEA scores of Myc-Targets-V2 between low and high ARID1A groups (adjusted *p* value =0.0013)**. (C)** Network visualisation of Myc-targets leading-edge gene expression in low and high ARID1A. Pearson correlation performed to assess negative and positive relationships between genes in the low and high ARID1A expressing groups. Node size represents the average gene expression and edge thickness represents correlation strength (|r| ≥ 0.7). **(D)** Representative images of c-Myc, CDK4 and Ki67 staining with box plots of percentages assessed in high vs low ARID1A groups (n=234). *p* values ≤0.05 were considered statistically significant.

### Transcription factor signatures in high ARID1A group suggest a shift toward transcription activation and regenerative stem cells signatures

Other transcriptional differences between the low and high ARID1A groups were explored using TF enrichment analysis (Figure 5A). The low ARID1A group was characterised by enrichment of HNF1B, E2F6 and MXI1, transcription factors associated with adenoma biology, cell-cycle regulation and tumour-suppressive functions (28–30). STRING analysis revealed that these TFs were predominantly involved in stem-cell maintenance, transcriptional repression and positive regulation of differentiation, consistent with their annotation as transcriptional repressors by UniProt keyword enrichment (Supplementary Figure 3, panel A).

**Figure 5.**
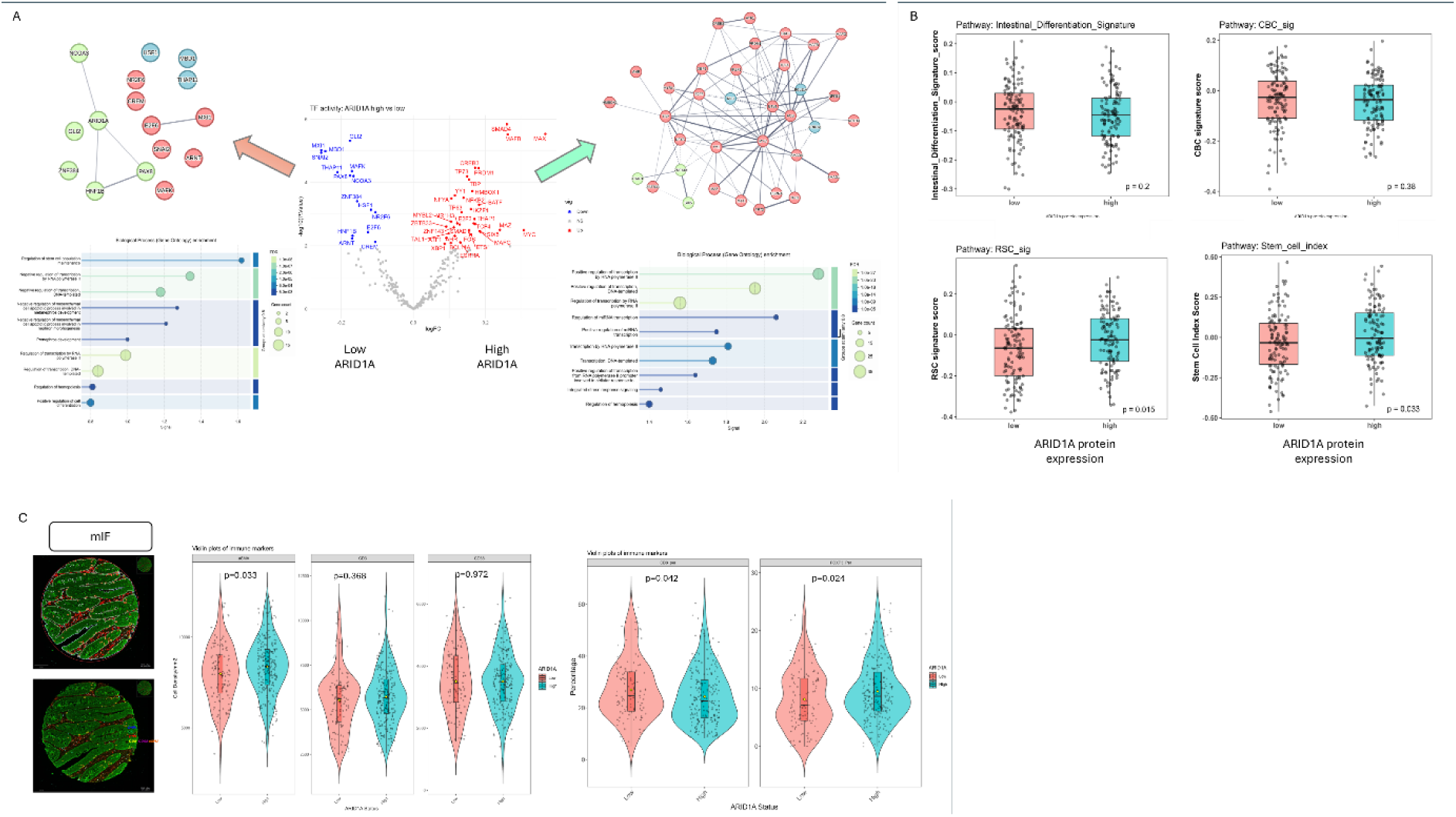
Transcription factors, stem cell and immune cell profiles in pre-cancerous polyps in relation to ARID1A expression. **(A)** Volcano plot showing differentially expressed TF using DoRothEA database with STRING network analysis performed for significant differentially expressed TFs in low and high ARID1A groups. ARID1A gene was added to the list of genes to elucidate TF with direct relationship with ARID1A **(B)** ssGSEA signature scores of intestinal, CBC and RSC stem cells and stem cells index in low and high ARID1A groups. **(C)** Representative mIF staining of immune cells using CD3, CD8, FOXP3, CD68, aSMA and pan-cytokeratin with violin plots showing cell densities of aSMA^+^, CD3^+^ and CD68^+^ cells and percentages of CD3^+ve^CD8^+ve^ and CD3^+ve^FOXP3^+ve^ cells in low vs high ARID1A groups. *p* values ≤0.05 were considered statistically significant.

In contrast, the high ARID1A group was enriched for TFs associated with transcriptional activation, including TBP, YY1, MAX, MYC and TP63 (Figure 5-panel A), which have established roles in angiogenesis, stem-cell function and cancer development (31–34). These factors were similarly annotated as transcriptional activators (Supplementary Figure 3, panel B).

Given these transcriptional differences, we next examined intestinal stem-cell programs. While overall intestinal stem-cell signatures did not differ between ARID1A groups, the regenerative stem-cell (RSC) signature and stem-cell index (35) were significantly enriched in the high ARID1A group (*p*=0.015 and *p*=0.033; respectively), whereas the crypt base columnar (CBC) signature remained unchanged (Figure 5-panel B). These findings suggest a potential association between high ARID1A expression and a regenerative stem-cell program, rather than widespread alterations in stem-cell activity, although the biological significance of this selective RSC enrichment requires further investigation.

### Transcriptomic and cellular evidence of immunosuppressive microenvironment in relation to ARID1A expression in pre-malignant colonic polyps

We performed mIF staining to assess immune cell infiltration (Figure 5-panel C). While no significant differences were observed in the abundance of CD3⁺ T cells or CD68⁺ macrophages between the groups, the differences in αSMA⁺-stromal cells and CD3 subsets (CD8 and FOXP3) were significant (*p*=0.033, *p*=0.042 and *p*=0.024, respectively; Figure 5- panel C. Tissues with high ARID1A expression exhibited significantly lower levels of CD8⁺ cytotoxic T cells and higher proportions of FOXP3⁺ regulatory T cells compared to the low ARID1A group. Together, these shifts in stromal and immune composition coupled with epithelial-intrinsic dysregulation point toward coordinated remodelling of the early polyp niche that may prime high-ARID1A tissues for immune evasion and possibly accelerate progression.

## Discussion

This study demonstrated that ARID1A protein expression identifies biologically distinct pre- malignant colorectal lesions with differing risks of metachronous polyp development. Although mutational profiling confirmed frequent alterations in canonical colorectal cancer drivers (including *APC*, *KRAS*, and *TP53*), *ARID1A* mutational status alone did not predict future polyp occurrence. In contrast, ARID1A expression was independently associated with metachronous disease, suggesting that protein-level assessment captures functional biological states not reflected by genetic sequencing alone. Importantly, the predictive value of ARID1A was most evident in lesions classified as low-risk using conventional pathological criteria, indicating that ARID1A can refine current risk stratification approaches and identify patients who could benefit from more intensive surveillance despite apparently favourable index criteria.

These findings extend previous reports demonstrating context-dependent roles of ARID1A in colorectal tumorigenesis. ARID1A loss has been associated with the elimination of chromosomally unstable cells and reduced tumour initiation in certain experimental models (9, 13). Our findings suggest that this effect may persist in established adenomas, as lesions with low ARID1A expression were associated with a reduced risk of subsequent metachronous polyps. Conversely, adenomas retaining high ARID1A expression may represent a biologically distinct state in which preserved chromatin-remodelling capacity supports sustained activation of oncogenic transcriptional programs and enhanced epithelial fitness. This interpretation is consistent with established mechanistic models of intestinal tumorigenesis, in which early lesions that maintain strong proliferative and survival advantages are more likely to undergo continued clonal expansion.

While ARID1A is traditionally characterised as a tumour suppressor in advanced malignancies, functional genetic models in the mouse colon have demonstrated that an intact SWI/SNF complex is required for conventional, *APC*-driven intestinal stem cell signalling and renewal

(36). Specifically, knock-out models showed that *APC*-mutant mice rapidly develop widespread colonic adenomas (9), however, concurrent ablation of *Arid1a* paradoxically suppresses this tumour burden in murine models (8, 12), demonstrating that an intact SWI/SNF complex is required to facilitate downstream Wnt signalling and proliferation programs.

Transcriptomic and transcription factor profiling provided clear biological explanations for these clinical observations, revealing contrasting regulatory landscapes between low and high ARID1A expression levels. ARID1A-low lesions were characterised by the enrichment of transcriptional repressors involved in differentiation, suppression of Myc activity, cell-cycle restraint and maintenance of tissue homeostasis. In contrast, high-ARID1A polyps demonstrated activation of Myc-driven transcriptional programs, a well-established driver of colorectal cancer proliferation downstream of *APC* mutations (37, 38) alongside a regenerative stem-cell signature. This hyper-proliferative tissue tone suggests that ARID1A- high polyps exist in a biologically permissive state that suppresses mechanisms promoting cell cycle exit, favouring continued epithelial renewal and progenitor-like behaviour (36).This shift toward a stem-like state has been previously shown to facilitate tumour plasticity (35), therapy resistance (39) and metastasis to the liver in colorectal cancer (40). These observations support the hypothesis that ARID1A expression influences early neoplastic evolution through its effects on intrinsic epithelial processes.

At the network level, TF profiling revealed contrasting regulatory landscapes between low and high ARID1A expression levelsin polyps. ARID1A-low tissues were enriched for repressors that maintain differentiation and antagonise Myc activity. Amongst these TFs, THAP11, a regulator of the cell cycle via negative regulation of c-Myc expression (41), and MXI1 are known to inhibit c-Myc transcription via dimerisation with Max protein and inhibition of the promoter region of Myc gene. MXI1 expression is lost in colorectal cancer suggesting a role as an antagonist of Myc gene (42). The expression of E2F6 in the low-ARID1A group may contribute to blocking cells undergoing G1/S cell cycle progression (29). MBD-1 (methyl- CpG-binding domain protein 1) is a repressor of transcription via DNA methylation control (reviewed in (43)). NR2F6 is involved in controlling the immune response and protecting the gut barrier homeostasis (44). Altogether, the TF profiles in the low-ARID1A group suggest that regulatory mechanisms are involved in the suppression of proliferation and promotion of cellular differentiation. In contrast, ARID1A-high polyps express activators linked to proliferation, angiogenesis, and stemness. PAX8 expression has been reported to be absent in normal epithelial colonic cells and is upregulated in cancer tissues (45), whereas HNF1B is high in normal and low-grade adenoma and completely lost in colorectal adenocarcinoma tissues suggesting a role as a tumour suppressor in colon tissues (28). Myc is a key gene that connects multiple TFs in the STRING network. Other enriched genes included TAL1 and BCL11A genes which are involved in haematopoiesis and stemness as well as E2F3 which regulates S-phase proliferation. THAP1, which is implicated in DNA repair, was also enriched, suggesting the potential accumulation of DNA damage during polyp progression (46–48).

Beyond these intrinsic epithelial programs, ARID1A expression levels appeared to clearly organise the surrounding microenvironmental niche. ARID1A-high polyps exhibited a significant increase in *α*SMA-positive stromal cells, suggesting a fibroblast-rich niche that may actively support epithelial stemness in a manner similar to intestinal subepithelial myofibroblasts (ISEMFs) (49). Furthermore, while total immune infiltration was comparable between the groups, high-ARID1A tissues demonstrated a marked compositional shift toward an immunosuppressive microenvironment, signified by an enrichment of FOXP3+ regulatory T-cells and a relative depletion of CD8+ cytotoxic T-cells. This shift is a recognised mechanism of immune evasion and poor prognosis in CRC (50, 51). In contrast to this observation, a previous study proposed that patches of ARID1A loss in the normal colon can induce an immunosuppressive neighbouring environment, however the number of patients included was limited (n=18 specimens) leading to no statistical significance upon analysis (8).

Together, these findings suggest that ARID1A is not only a predictive marker for metachronous polyp risk but also a key regulator of epithelial dynamics, stromal remodelling and immune evasion during early colorectal tumorigenesis. Incorporating ARID1A assessment into routine pathology could transform the current surveillance standards and improve patient outcomes.

### Strengths and Limitations

The major strength of this study lies in its multi-omic integration, bridging histopathological, mutational, transcriptomic and microenvironmental protein-expression data within a large, well-characterized clinical screening cohort. Furthermore, the use of clinically relevant outcomes, namely metachronous polyp occurrence, enhances the translational relevance of this study.

This study had several limitations. First, TMAs are primarily used as the evaluation platform and may not fully capture intralesional heterogeneity. Further validation is required for whole tissue sections from different demographic cohorts. Second, the synchronous lesions were not evaluated. Therefore, it remains uncertain whether the biology of the index polyp accurately reflects the entire colonic field or whether it applies equally across all simultaneous polyp sizes and subtypes. Finally, our mutational analysis was restricted to a targeted gene panel. Broader genomic, epigenomic, and methylation profiling may identify additional cooperative pathways underlying metachronous colorectal lesion risk.

### Clinical Implications and Future Directions

From a clinical perspective, these findings suggest that incorporation of ARID1A immunohistochemical assessment into routine pathological evaluation could improve risk stratification after polypectomy and potentially inform personalised surveillance strategies especially in low risk groups with fewer index polyps, low grade dysplasia and from the left colon. Given that ARID1A staining can be readily implemented using standard pathology workflows, its translation into clinical practice may be feasible if prospectively validated. Future studies should focus on validating these findings in larger independent cohorts, assessing ARID1A expression across synchronous and metachronous lesions, and defining the mechanistic relationship between ARID1A, fibroblast-rich stromal niches, stem-cell programmes, and immune regulation. Elucidating whether ARID1A directly drives these biological processes or serves as a marker for a broader high-risk state remains an important unanswered question. Such studies may ultimately identify novel preventive strategies aimed at interrupting early colorectal neoplastic progression.

## Declarations

### Ethical approval

Ethical approval was granted through the West of Scotland Research Ethics Committee (GSH/20/CO/002).

### Data availability statement

The clinical database is accessible through the NHS SafeHaven Trusted Research Environment (TRE) subject to appropriate ethical approval and completion of mandatory information governance training modules. Alternatively, access can be arranged by contacting Professor Joanne Edwards. Genomics data can be accessed through Datacite DOIs:10.5525/gla.researchdata.1223, 10.5525/gla.researchdata.1498 and 10.5525/gla.researchdata.1544. Transcriptomic datasets generated and/or analysed during the current study are available upon reasonable request from the INCISE project director, Professor Joanne Edwards.

### Patient and public involvement (PPI)

Patient involvement in this study was facilitated through a dedicated meeting with the INCISE patient steering group https://www.gla.ac.uk/research/az/translationalcancerpathology/forpatientsandthepublic/.

During this session, the study data were presented and discussed. The group feedback helped inform the research questions explored in this study and contributed to shaping ideas for future research.

## Acknowledgment

We would like to acknowledge the Glasow Tissue Research Facility (GTRF), Glasgow University, with extended thanks to Dr Hannah Morgan for the TMA construction, sectioning and slide scanning. We would also like to thank the Greater Glasgow and Clyde Biorepository for providing polyp tissue samples essential for this study. We also acknowledge Professor John Le Quesne’s group at the CRUK Scotland Institute Deep Phenotyping Advanced Technology Facility (RRID:SCR_027366), with a special acknowledgment for Leah Officer-Jones, Silvia Martinelli, Rachel Baird, and Fiona Ballantyne for assistance with scanning mIF images and with channel unmixing on the InForm software. Finally, we would like to express our gratitude to the INCISE patient steering group for supporting this research.

## Authors’ contributions

**AA** database curation, study design, data analysis and manuscript drafting**, AM** ARID1A scoring, data quality control and manuscript editing; **KKA** staining of whole tissue sections; **SM** and **MH** staining and scoring ARID1A in full sections; **AW** genomics data processing; **NF** transcriptomics data processing/analysis and manuscript editing; **NM** pathology review; **GPL** resource management and manuscript revision; **JH** tissue and pathology database management; **TI** multiplex IF image analysis, critical reading and editing of manuscript; **CWS** manuscript revision and critical feedback for data analysis; **MJ** INCISE patient cohort clinical data collection; **PDD** Supervision of transcriptomics data management; **STM** Supervision of INCISE patient cohort clinical data collection and analysis; **JE** study supervision, study design and manuscript revision.

## Figure legends (supplementary)

**Supplementary Figure 1.**
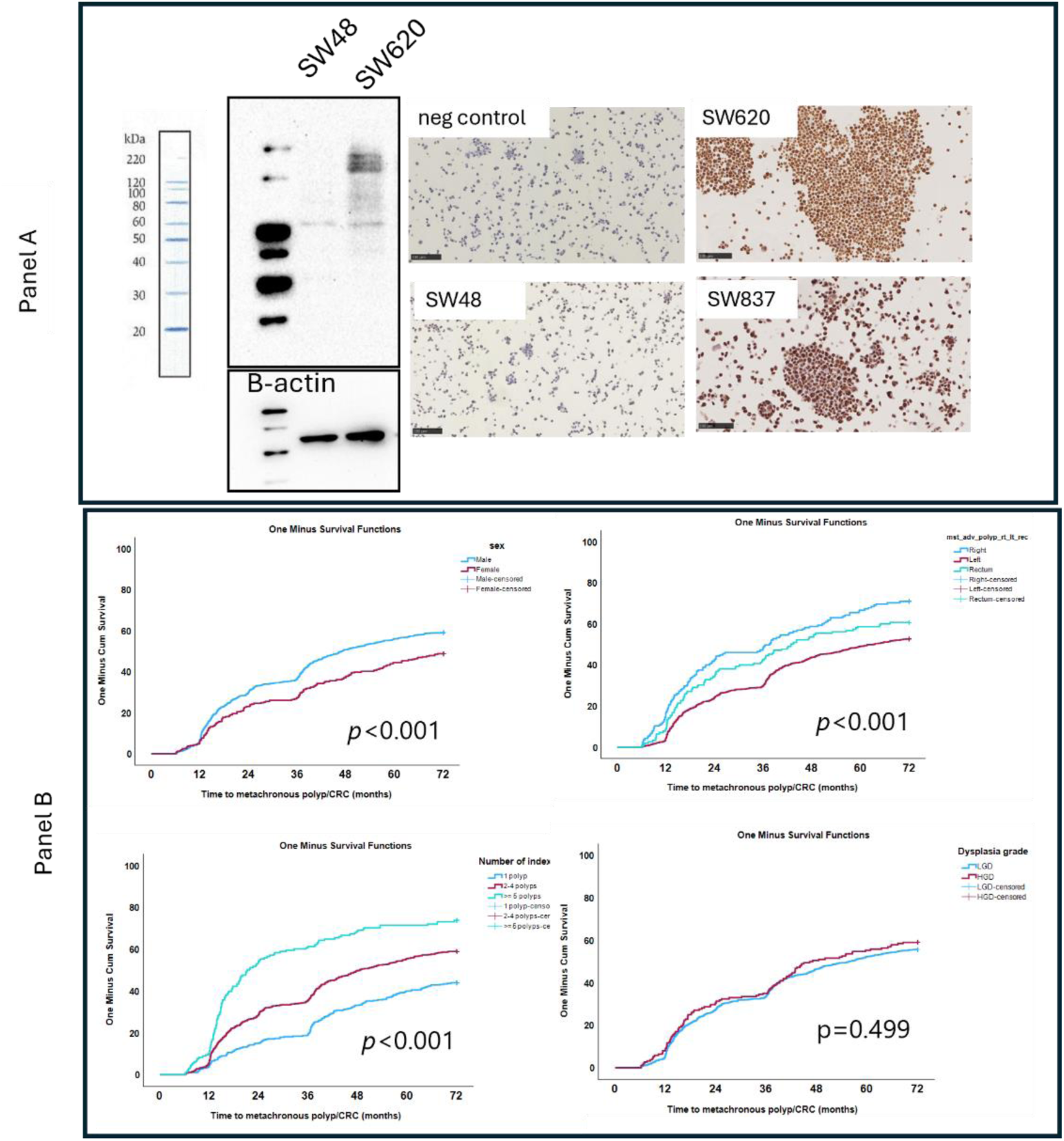
**(panel A)** ARID1A Western blotting and IHC staining in ARID1A-mutant SW48 vs wildtype SW620 and SW837 confirmed the specificity of the ARID1A antibody; ARID1A molecular weight is ∼220 kDa. Another band of truncated form of ARID1A appeared at ∼65 kDa. **(panel B)** Survival analysis and Log-rank test results of confounding factors predicting risk of metachronous colorectal lesions including sex (*p*<0.001), location of index polyps (*p*<0.001), number of polyps (*p*<0.001) and dysplasia grade (*p*=0.499). *p* values ≤0.05 were considered statistically significant.

**Supplementary Figure 2.**
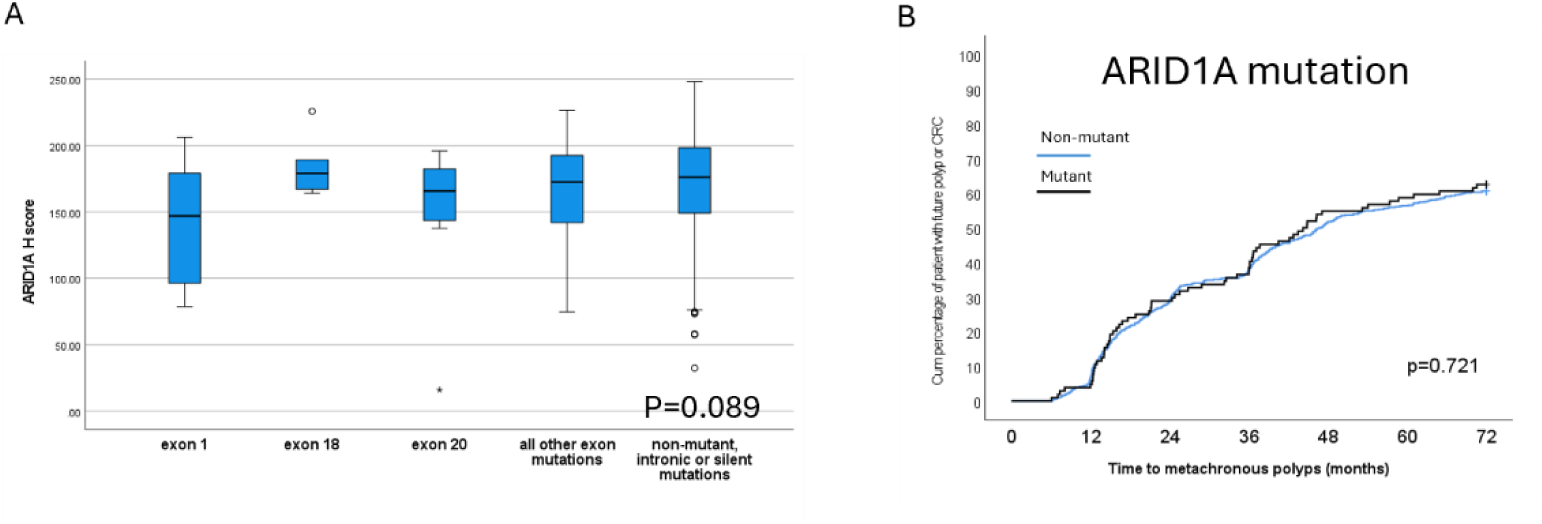
Supplementary figures for mutational data in Figure 3. **(A)** ARID1A protein expression based on mutation types showing no significant differences in ANOVA analysis (*p*=0.089. (B). Kaplan-Meier curve and survival analysis of the predictive significance of *ARID1A* gene mutation in terms of metachronous polyp development showing no significant association with metachronous lesions (*p*=0.721).

**Supplementary Figure 3.**
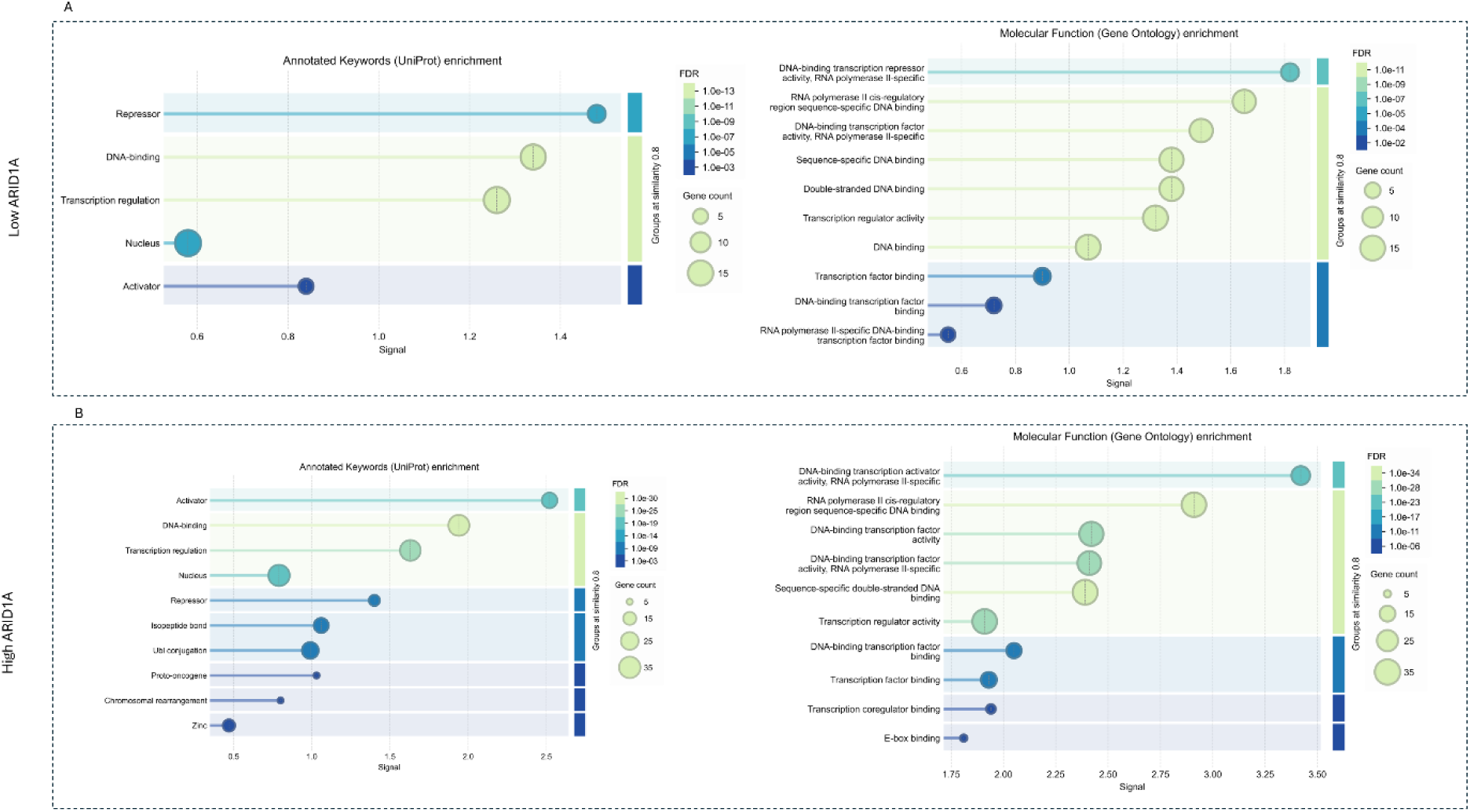
STRING analysis of transcription factors enriched in low or high ARID1A expression showing pathways. Panel A and B show the Annotated keywords Uniprot and Gene-ontology molecular functions pathways enrichment in the low and high ARID1A expression groups; respectively. Low ARID1A group was marked by repression of transcription whereas high ARID1A expression group showed high representation of activators and transcription acceleration pathways.

**Supplementary table 1.**
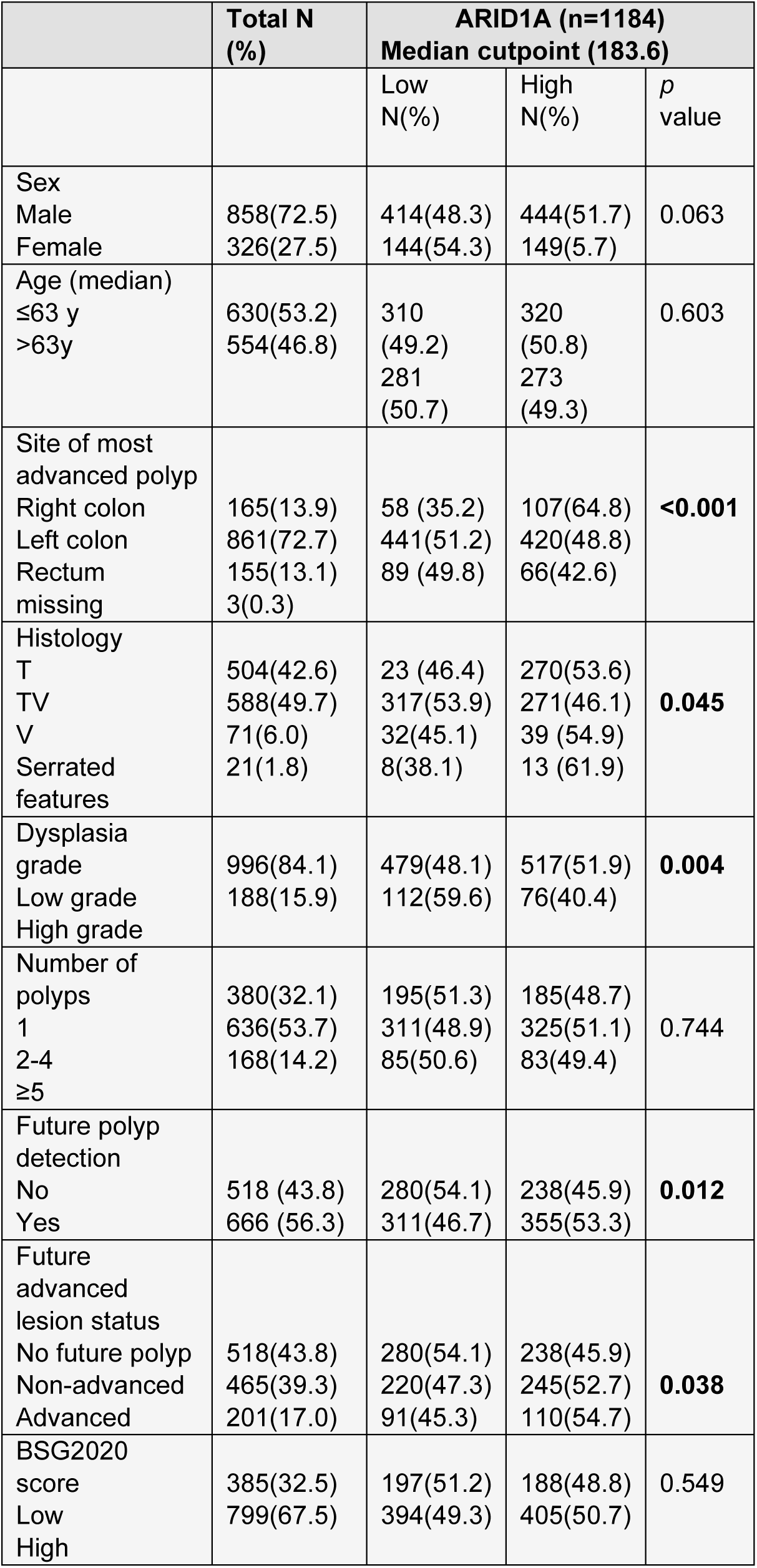
Chi-squared test results of epithelial ARID1A histoscores with clinicopathological criteria using the median as a cutpoint.

## Supplementary Methods

### Western blotting

Based on information in DepMap portal (https://depmap.org/portal), SW48 cell line harbours a deletion mutation in ARID1A whereas SW620 and SW837 are wildtype. Cell line authentication was confirmed using Short Tandem Repeat (STR) profiling assay. Cell lysates were prepared from SW48, SW620 and SW837 using RIPA buffer supplemented with protease and phosphatase inhibitors. Pellets (∼1.5 × 10⁶ cells) were lysed in 150μL buffer via repeated syringe shearing, incubated on ice, and centrifuged at 14000 g for 5 min to collect supernatants. Protein concentrations were quantified using the Bradford assay with bovine serum albumin (BSA) standards. Lysate concentrations were normalised to 1 mg/mL, mixed with 4X loading dye (Nu-Page, Invitrogen) supplemented with beta-mercaptoethanol, boiled for 5 min and stored at −20°C until use. 20 ug of individual protein lysates were separated via Bio-Rad Mini-Protean TGX 4-20% pre-cast gels (#4561093, BioRad), transferred to nitrocellulose membranes, and verified with Ponceau S staining to ensure successful transfer. Membranes were blocked with 5% BSA for 1h at room temperature (RT), then incubated with primary antibodies for ARID1A (Cell Signaling, cat#12354, 1:1000 at 4°C overnight) and beta-actin (loading control, 1:5000, Cell Signalling # 8H10D10, 1 h at RT) followed by appropriate HRP-conjugated secondary antibodies for 1h at RT. Detection was carried out using ECL substrate (#2106, Thermofisher) and visualised using a BioRad Chemi-Doc system.

### Supplementary methods for IHC

Cell pellets prepared from SW620, SW48 and SW 837 colorectal cancer cell lines were fixed in 10% formalin, dehydrated in ethanol and xylene, embedded in paraffin and sectioned.

FFPE sections from TMA (2.5-μm sections), and cell pellets or whole tissue sections (4 μm) were used. Cell pellets, TMA and full section slides were de-waxed and antigen retrieved using a PT-module (Agilent) and dewax-antigen retrieval (Epredia). Ultravision Quanto detection system (Epredia) was used as follows: endogenous peroxidase was blocked with 3% H2O2 for 10 min followed by a protein blocking step (8 minutes). Primary antibody step using ARID1A (Cell Signaling cat#12354; 1:1500, overnight at 4 °C) CDK4 (Cell Sigaling #D9G3E, 1:100, 1 hour at room temperature (RT)) or Ki67 (Agilent, #M7240 1:100 for 30 min at RT) was applied. The tissues were treated with an amplifier for 10 min followed by an HRP polymer step (10 min). The signal was visualised with 3,3′-diaminobenzidine and counterstained with haematoxylin Gill III formula (Leica Biosystems), dehydrated in ethanol and xylene followed by mounting with ClearVue (Epredia). The c-Myc antibody (Abcam #ab32072, 1:800 for 35 min) was stained using the Leica Bond Rx autostainer and Refine Kit (Leica Biosystems). Slides were scanned at 20x magnification using a Hamamatsu S60 NanoZoomer (Hamamatsu, Japan).

### Multiplex IF staining (mIF)

TMA sections of 2.5-µm thickness were stained using the Link 48 autostainer (DAKO) and UVQ kit (Epredia). The primary antibody incubation was performed for 30 minutes at room temperature. The UVQ System HRP (Epredia) was used for blocking, signal amplification, and secondary antibody incubation as described in the IHC protocol. Opal-TSA fluorophores (Akoya Biosciences, Marlborough, MA, USA) were applied at a 1:200 dilution for 10 minutes. Following each round of antibody staining, an additional antigen retrieval step was performed using the PT module and Dewax-HIER-H (Epredia). Nuclear counterstaining was performed with Spectral DAPI (1:1000 dilution, AKOYA) for 5 minutes, and slides were mounted using ProLong Diamond Antifade Mountant (ThermoFisher Scientific).

### List of antibodies used in mIF

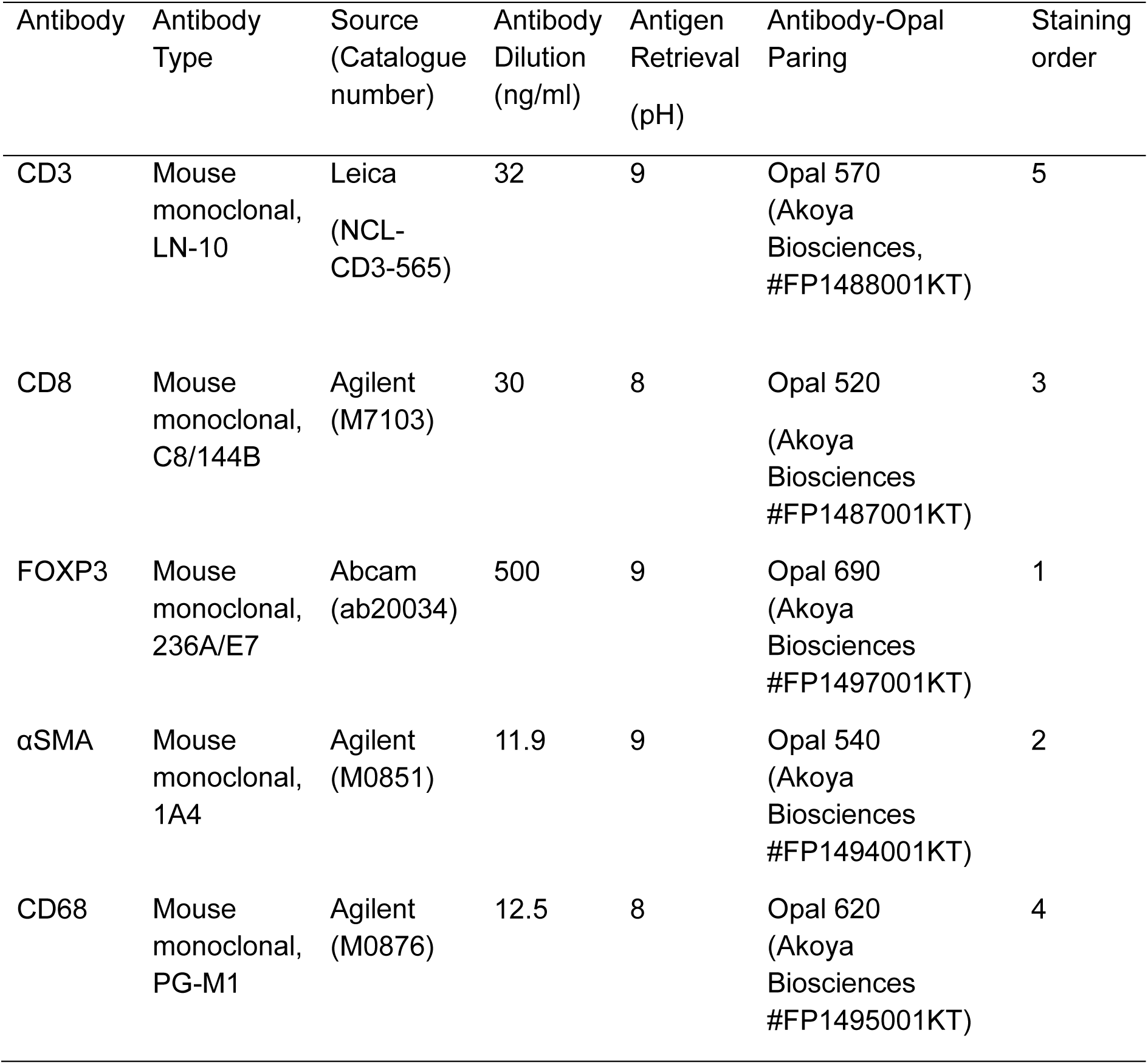

| Antibody | Antibody Type | Source (Catalogue number) | Antibody Dilution (ng/ml) | Antigen Retrieval (pH) | Antibody-Opal Paring | Staining order |
| --- | --- | --- | --- | --- | --- | --- |
| CD3 | Mouse monoclonal, LN-10 | Leica (NCL-CD3-565) | 32 | 9 | Opal 570 (Akoya Biosciences, #FP1488001KT) | 5 |
| CD8 | Mouse monoclonal, C8/144B | Agilent (M7103) | 30 | 8 | Opal 520 (Akoya Biosciences #FP1487001KT) | 3 |
| FOXP3 | Mouse monoclonal, 236A/E7 | Abcam (ab20034) | 500 | 9 | Opal 690 (Akoya Biosciences #FP1497001KT) | 1 |
| αSMA | Mouse monoclonal, 1A4 | Agilent (M0851) | 11.9 | 9 | Opal 540 (Akoya Biosciences #FP1494001KT) | 2 |
| CD68 | Mouse monoclonal, PG-M1 | Agilent (M0876) | 12.5 | 8 | Opal 620 (Akoya Biosciences #FP1495001KT) | 4 |

Slides were scanned using a PhenoImager-HT (Akoya Biosciences) and, underwent spectral unmixing by InForm using a library of single stained slides.

### Analysis of IHC images

QuPath (v0.5.0) was used to analyse the IHC images. Positive cell detection was used to identify high, moderate and low expression of ARID1A and Histoscores (H-scores; 0-300) were calculated. An object classifier using machine learning Random Trees classification segmented epithelial and lamina propria cells. For c-Myc, CDK4 and Ki67, a single threshold was used to detect positive cell within the epithelial cells. Cores with >100 adenoma cells were included in the analysis. 10% of scores were validated manually.

### Analysis of mIF images

Images were analysed using the QuPath (Version 0.5.1). Tissues and staining artefacts were visually inspected and excluded from regions of interest (ROI). Tissue segmentation to distinguish the epithelium and lamina propria was performed by a pixel classifier generated by the Random Trees algorithm using pan-cytokeratin, α-SMA, and DAPI as detection channels. Object classifiers for CD3^+^, CD8^+^, and FOXP3^+^ cells were generated based on CD3 channel for detection and using intensity threshold. Object classifiers were applied sequentially to detect CD3^+^, CD3^+^CD8^+^, and CD3^+^FOXP3^+^ cells. CD68^+^ and α-SMA^+^ cells were identified by cell detection using each channel. Cell density (cells/mm^2^) was calculated using the number of detections divided by area of the ROIs.

### Supplementary methods for DNA extraction, sequencing and somatic mutational analysis

DNA was extracted using the Maxwell RSC instrument (Promega) using two 10µm whole tissue section curls and assessed for quality using Qubit and TapeStation. Variant calling was performed.by the Genomic Innovation Alliance (Glasgow, UK). Samples with low DNA concentration (<10 ng/µL) or poor integrity (DIN <3.0) were excluded. Libraries were prepared using the Agilent SureSelect XT2 HS2 protocol with Cancer Plus panel enrichment and sequenced on an Illumina NovaSeq 6000, aiming for 500× mean coverage. This next-generation sequencing (NGS) data processing was performed using HOLMES pipeline v1.3.1. FASTQ files were aligned to hg38 with BWA v0.7.15, SNVs and indels were called using deepSNV/Shearwater and Pindel, structural variants with Brass, and copy number alterations with geneCN. Variants were annotated with CAVA. Variants underwent quality filtering based on allele frequency, read depth, mapping quality and presence in public databases. Sequencing QC thresholds included: (1) mean coding depth >50×, (2) insert size >120 bp, (3) non-reference read rate <3.5%, (4) discordant pairs <10% and (5) on-target reads >20%. Samples were analysed in tumour-only mode using a panel of normals from 21 healthy tonsil samples (unmatched normal) to filter artifacts.

### Supplementary methods for Transcriptomics analysis

Quality control and preprocessing included the following steps. Probe variance across all samples was calculated using the *sapply* function. Probes with a variance less than the 25^th^ percentile were removed (n = 5640 probes). Samples with a low read count (< 95% confidence interval [<2.28×10^6^ counts]) were identified (*stats* v4.2.3) and removed. Samples with incomplete batch information were excluded (n =5). The counts were batch corrected using *ComBat_seq* (sva v3.46.0). Probe IDs were mapped to gene symbols with duplicated genes collapsed using *MaxMeans* (*WGCNA* v1.72-1), resulting in 14,993 genes. Following outlier review n=20 samples were removed from the INCISE cohort (4). Raw counts were normalised with DESeq2. Group-wise enrichment was performed using fgsea with MSigDB Hallmark gene sets in R. Individual pathway scores (10–500 genes) from MSigDB including Hallmarks) or previously reported intestinal stem cell (23), crypt-base columnar (CBC) and regenerative stem cell (RSC) signatures [21,22] were computed using ssGSVA. Pearson correlation analysis within low and high ARID1A groups was used to identify gene-gene relationships within the leading-edge genes. Transcription factor (TF) activity was inferred using DoRothEA (levels A-C) with the viper algorithm to compute regulon activity scores. Differential TF activity between ARID1A groups was assessed using linear modelling with empirical Bayes moderation (limma).

In this study, a selected group of patients with bulk transcriptomic data was used (n=234) based on the upper and bottom 10% percentile of ARID1A weighted histoscores.

